# The Influence of Intimate Partner Violence on Non-Facility Births Attended by Unskilled Providers in Tanzania: Evidence from the 2022 Demographic and Health Survey

**DOI:** 10.1101/2025.03.28.25324825

**Authors:** Fabiola Vincent Moshi, Keiko Nakamura, Yuri Tashiro, Ayano Miyashita, Hideko Sato, Mayumi Ohnishi

## Abstract

**Background:** Non-facility births assisted by unskilled attendants remain a significant public health concern in Tanzania. Despite government efforts to expand access to maternal health services, such as increasing the number of healthcare facilities, scaling up skilled birth attendants, and eliminating financial barriers, many women continue to give birth under the care of unskilled attendants. This study aimed to assess the influence of intimate partner violence (IPV) as a predictor of non-facility births attended by unskilled individuals in Tanzania.

**Method:** The study used the 2022 Tanzania Demographic and Health Survey and Malaria Indicator Survey (2022 TDHS-MIS) dataset, with a weighted sample of 2,284 women of reproductive age who gave birth two years preceding the survey and were included in intimate partners violence survey. Descriptive analysis determined the magnitude of both experiences in different forms of intimate partner violence and non-facility. Both univariate and multiple regression analyses were used to identify significant determinants of non-facility birth.

**Result:** A total of 438 women (19.2%) had non-facility births assisted by unskilled attendants. After adjusting for confounding factors, key predictors of non-facility births included ever experiencing any form of IPV (aOR=1.383, p=0.007). Other significant factors were educational attainment, with women having no formal education (aOR=2.718, p<0.001) and primary education (aOR=1.785, p=0.011) being more likely to give birth outside a facility; wealth index [poorest (aOR=7.468, p<0.001), poorer (aOR=4.434, p<0.001), middle (aOR=3.27, p=0.001) and richer (aOR=2.734, p=0.005)]; Antenatal Care (ANC) attendance less than four visits (aOR=1.574, p<0.001); number of children ever born [Two to four (aOR=2.408, p<0.001), more than four (aOR=3.119, p<0.001)] and ever terminated pregnancy (aOR=1.525, p=0.026).

**Conclusion:** Women who experienced IPV had significantly higher odds of non-facility births. Other crucial predictors included low educational attainment, poverty, having multiple children, attending fewer than four ANC visits, and a history of pregnancy termination. Since IPV is a modifiable risk factor, early detection during ANC visits and the implementation of targeted interventions could greatly enhance access to facility-based births with skilled professionals, thereby improving maternal and child health outcomes.

## Background Information

Maternal mortality remains disproportionately high in low- and middle-income countries, where inadequate access to skilled birth attendants and facility-based deliveries worsens the situation.(1). In 2020, sub-Saharan Africa had 545 maternal deaths per 100,000 live births as compared to 4 in Australia and New Zealand (1). Skilled health professionals, as defined by Sustainable Development Goal (SDG) indicator 3.1.2, are trained maternal and newborn care specialists who meet national and international education, training, and regulatory standards (2). They possess the expertise to deliver and advocate for evidence-based, human-rights-centered, high-quality, and culturally responsive care that upholds the dignity of women and newborns (2). Their role encompasses supporting the natural physiological processes of labor and childbirth, ensuring a safe and positive birthing experience, and promptly identifying, managing, or referring cases of maternal or neonatal complications to appropriate levels of care (2).

Globally, Skilled Birth Attendance (SBA) rates have increased from 61% in 2000 to 86% in 2023(3). Evidence underscores that progress in SBA is closely tied to rising household wealth, higher maternal education, urbanization, greater contraceptive knowledge, and critical markers of female empowerment (4). In Tanzania, access to skilled birth attendance remains disproportionately low in rural areas, with only 81% of women receiving SBA compared to 96% in urban settings. Moreover, having multiple children has been identified as a significant barrier, with just 73% of high-parity women accessing skilled birth attendance (5).

Tanzania has made substantial progress in improving maternal and child health by increasing the number of healthcare facilities, expanding the availability of skilled birth attendants, and eliminating financial barriers to maternal healthcare (6). Despite these efforts, non-facility births attended by unskilled individuals persist at significant rates, posing increased risks of maternal and neonatal morbidity and mortality. The Tanzanian health system has ensured the presence of skilled birth attendants across various healthcare facilities, including hospitals, health centers, dispensaries, and maternity homes. Births occurring outside these facilities are attended by untrained personnel. The absence of skilled care increases the risk of delayed recognition of labor complications, significantly heightening the likelihood of adverse birth outcomes.

While multiple socio-economic and health system-related factors contribute to the continued prevalence of non-facility births, the role of intimate partner violence (IPV) as a determinant of maternal health-seeking behavior remains underexplored. IPV is a pervasive issue in Tanzania, affecting a substantial proportion of women of reproductive age. It has been linked to adverse maternal health outcomes, including delayed or inadequate antenatal care (ANC) utilization, limited autonomy in decision-making regarding childbirth, and increased psychological distress, all of which may contribute to non-facility births. Understanding the relationship between IPV and non-facility births is essential for developing targeted interventions to improve maternal healthcare access and utilization.

This study aimed to assess the influence of IPV on the likelihood of non-facility births in Tanzania. Using data from the 2022 Tanzania Demographic and Health Survey and Malaria Indicator Survey 2022 Tanzania Demographic and Health Survey and Malaria Indicator Survey (TDHS-MIS). By identifying the extent to which IPV contributes to non-facility births, alongside other known determinants such as educational attainment, socioeconomic status, ANC attendance, and reproductive history, this study provides critical insights into the barriers preventing women from accessing skilled maternal healthcare.

## Method

### Study design

This study employed an analytical cross-sectional design, utilizing secondary data from the 2022 Tanzania Demographic and Health Survey and Malaria Indicator Survey (2022 TDH-MIS). The 2022 TDHS-MIS is a nationally representative survey that collected comprehensive information on various health-related indicators, including maternal and child health, reproductive health, and fertility.

### Study Population

The analysis for this paper focused on women of reproductive age (15-49 years) who gave birth within two years preceding the survey and were included in the intimate partners violence survey.

### Sample size

The 2022 TDHS-MIS utilized a stratified two-stage sample design to provide estimates for the entire country, including urban and rural areas in mainland Tanzania and Zanzibar. In the first stage, sampling points (clusters) were selected from enumeration areas delineated in the 2012 Tanzania Population and Housing Census (2012 PHC), with selection probability proportional to the size of each sampling stratum. A total of 629 clusters were chosen. In the second stage, 26 households were systematically selected from each cluster, resulting in a total of 16,312 households. All women aged 15–49 who were either usual residents or visitors in the household on the night before the survey interview were included in the 2022 TDHS-MIS and were eligible to be interviewed. Out of the 16,312 selected households, 15,907 were occupied. Of these, 15,705 households were successfully interviewed, yielding a response rate of 99%. Within the interviewed households, 15,699 women aged 15–49 were identified as eligible for individual interviews. Interviews were completed with 15,254 women, resulting in a response rate of 97%.

### Data extraction

We utilized the Individual Recode (TZIR82SV.ZIP) file to extract eligible participants from a total of 15,699 interviewed women, of whom 15,254 were of reproductive age. Two levels of extraction were conducted. Initially, we excluded women who did not give birth within the two years preceding the survey, resulting in a sample of 5,879 women. Subsequently, we excluded those who did not participate in the intimate partner violence survey, leaving us with 2,447 women. Among them, 163 did not respond to the question regarding the place of childbirth. Thus, the final sample used in this study comprised 2,284 women.

### Study variables

The outcome variable in this study was the place of childbirth, categorized into two groups: facility-based birth (coded as 0) and non-facility birth (coded as 1), with non-facility birth serving as the reference category. The primary explanatory variable was whether the individual had ever experienced IPV, which was dichotomized into either having experienced IPV or not. Four distinct types of IPV were assessed: emotional violence, less severe physical violence, severe physical violence, and sexual violence.

### Statistical Analyses

Data was analyzed using Statistical Package for Social Sciences (SPSS) version 25. We started by describing women’s background characteristics. A bivariate analysis with a chi-square test was performed to examine predictors of non-facility birth. A p-value less than 5% was considered significant.

### Ethical Consideration

The study utilized secondary data from the Demographic and Health Surveys (DHS), which is a publicly available dataset. In the case of Tanzania, the DHS data collection adhered to ethical standards set by the National Institute for Medical Research (NIMR) or an equivalent ethics committee. Prior approval was obtained for the study design and data collection, ensuring that ethical considerations were fully met in line with both local and international ethical guidelines

## Results

### Sociodemographic Characteristics

The majority of study participants were aged 20-34 years (71.3%), resided in rural areas of Tanzania (73.9%), had primary education (58.4%), had 2-4 children (55.6%), and had at least four antenatal care (ANC) visits (64.3%) Table 1

**Table 1:**
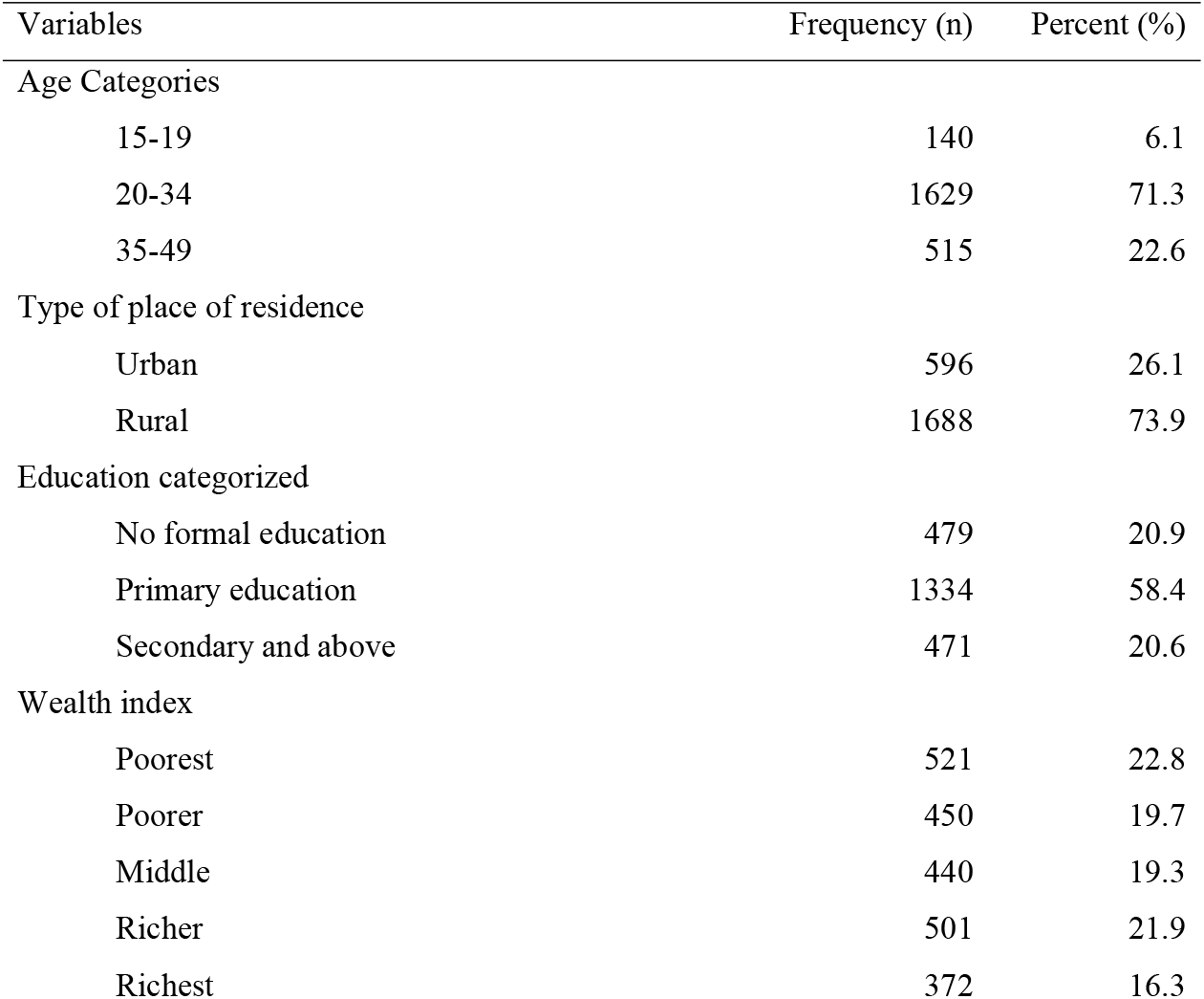
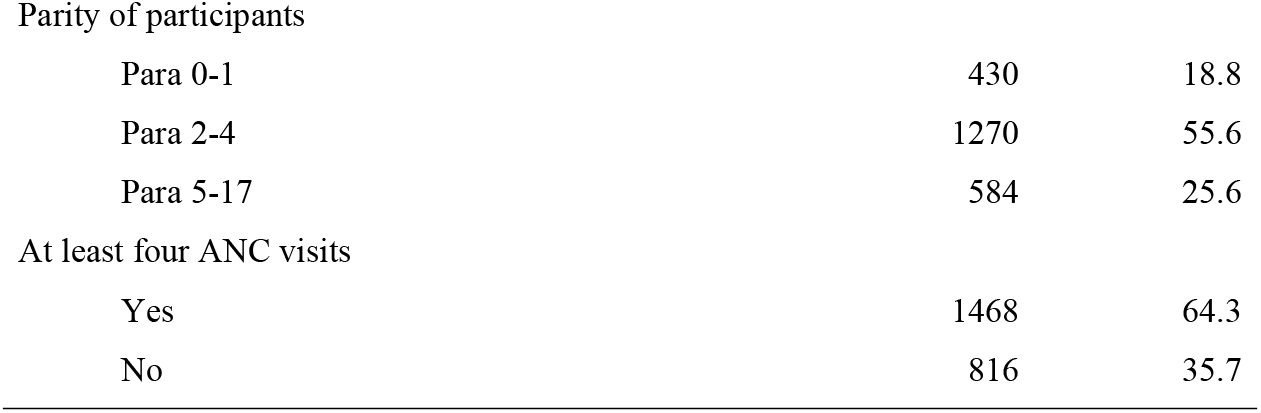
Sociodemographic characteristics.

| Variables | Frequency (n) | Percent (%) |
| --- | --- | --- |
| Age Categories |  |  |
| 15-19 | 140 | 6.1 |
| 20-34 | 1629 | 71.3 |
| 35-49 | 515 | 22.6 |
| Type of place of residence |  |  |
| Urban | 596 | 26.1 |
| Rural | 1688 | 73.9 |
| Education categorized |  |  |
| No formal education | 479 | 20.9 |
| Primary education | 1334 | 58.4 |
| Secondary and above | 471 | 20.6 |
| Wealth index |  |  |
| Poorest | 521 | 22.8 |
| Poorer | 450 | 19.7 |
| Middle | 440 | 19.3 |
| Richer | 501 | 21.9 |
| Richest | 372 | 16.3 |
| Parity of participants |  |  |
| Para 0-1 | 430 | 18.8 |
| Para 2-4 | 1270 | 55.6 |
| Para 5-17 | 584 | 25.6 |
| At least four ANC visits |  |  |
| Yes | 1468 | 64.3 |
| No | 816 | 35.7 |

### Intimate partner violence among women who gave birth in the last two years preceding the survey

Out of the surveyed women, 1314 (57.5%) reported experiencing control by their intimate partners, whereas 970 (42.5%) did not experience such control. Regarding emotional violence, 515 women (22.6%) experienced it, while 1769 (77.4%) did not. In terms of less severe physical violence, 610 women (26.7%) reported experiencing it, compared to 1674 (73.3%) who did not. Severe physical violence was experienced by 168 women (7.4%), while 2116 (92.6%) reported not experiencing it. Lastly, 166 women (7.3%) experienced sexual violence by their husbands or partners, whereas 2118 (92.7%) did not face any sexual violence (see Table 2 and Figure 1)

**Table 2:**
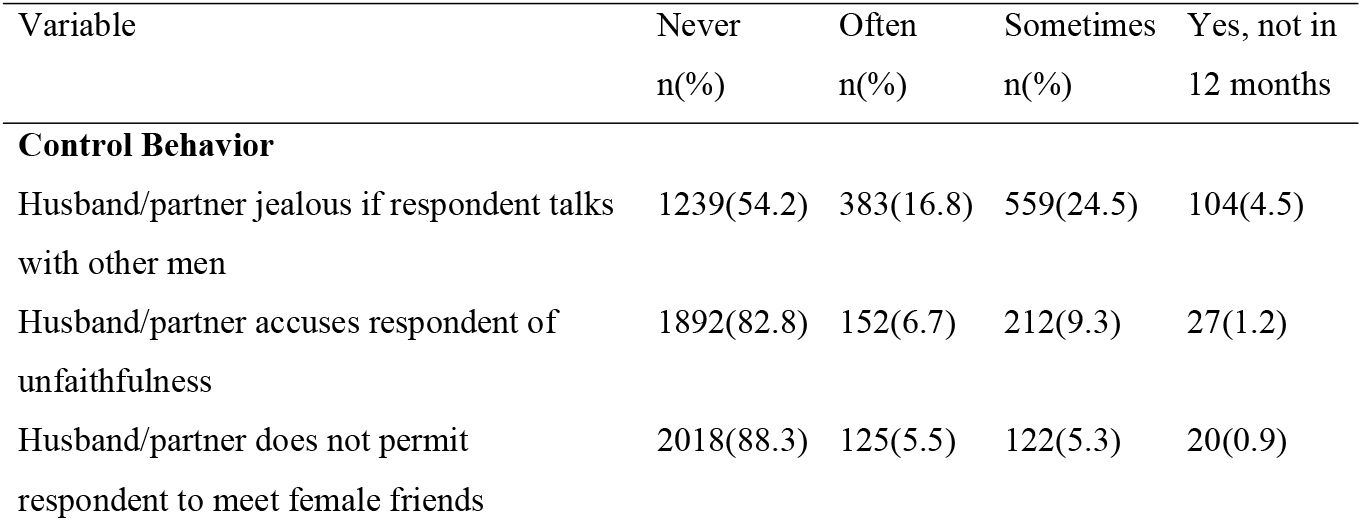
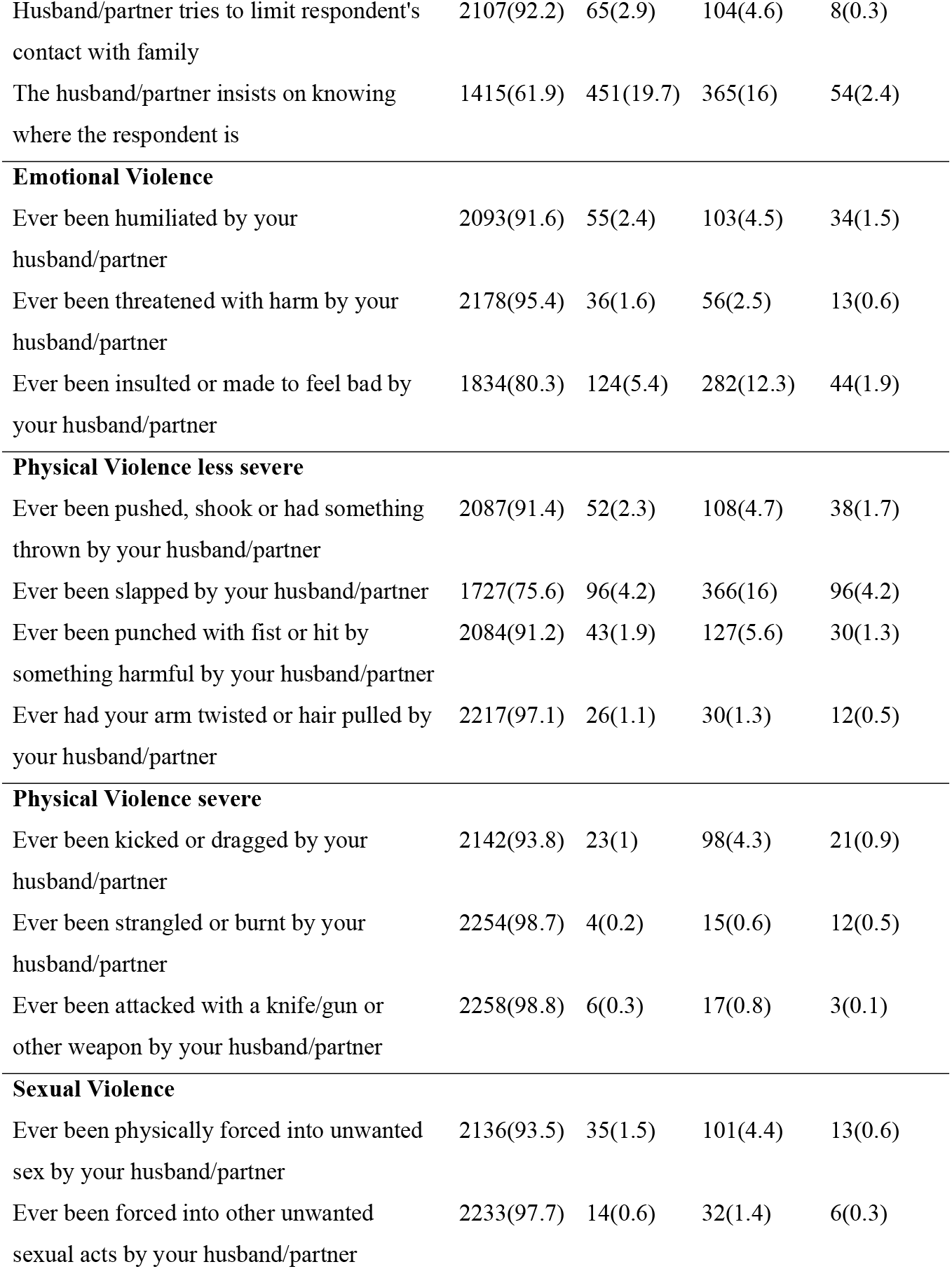
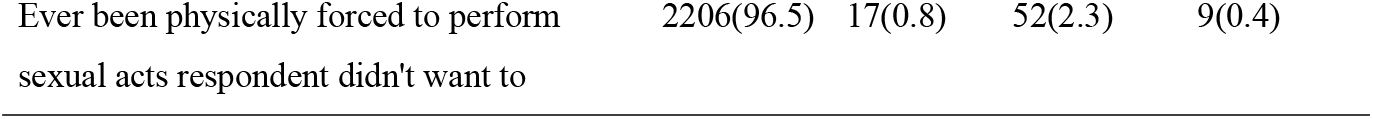
Intimate partner violence among women who gave birth in the last two years.

| Variable | Never<br>n(%) | Often<br>n(%) | Sometimes<br>n(%) | Yes, not in<br>12 months |
| --- | --- | --- | --- | --- |
| **Control Behavior** |  |  |  |  |
| Husband/partner jealous if respondent talks with other men | 1239(54.2) | 383(16.8) | 559(24.5) | 104(4.5) |
| Husband/partner accuses respondent of unfaithfulness | 1892(82.8) | 152(6.7) | 212(9.3) | 27(1.2) |
| Husband/partner does not permit respondent to meet female friends | 2018(88.3) | 125(5.5) | 122(5.3) | 20(0.9) |
| Husband/partner tries to limit respondent's contact with family | 2107(92.2) | 65(2.9) | 104(4.6) | 8(0.3) |
| The husband/partner insists on knowing where the respondent is | 1415(61.9) | 451(19.7) | 365(16) | 54(2.4) |
| **Emotional Violence** |  |  |  |  |
| Ever been humiliated by your husband/partner | 2093(91.6) | 55(2.4) | 103(4.5) | 34(1.5) |
| Ever been threatened with harm by your husband/partner | 2178(95.4) | 36(1.6) | 56(2.5) | 13(0.6) |
| Ever been insulted or made to feel bad by your husband/partner | 1834(80.3) | 124(5.4) | 282(12.3) | 44(1.9) |
| **Physical Violence less severe** |  |  |  |  |
| Ever been pushed, shook or had something thrown by your husband/partner | 2087(91.4) | 52(2.3) | 108(4.7) | 38(1.7) |
| Ever been slapped by your husband/partner | 1727(75.6) | 96(4.2) | 366(16) | 96(4.2) |
| Ever been punched with fist or hit by something harmful by your husband/partner | 2084(91.2) | 43(1.9) | 127(5.6) | 30(1.3) |
| Ever had your arm twisted or hair pulled by your husband/partner | 2217(97.1) | 26(1.1) | 30(1.3) | 12(0.5) |
| **Physical Violence severe** |  |  |  |  |
| Ever been kicked or dragged by your husband/partner | 2142(93.8) | 23(1) | 98(4.3) | 21(0.9) |
| Ever been strangled or burnt by your husband/partner | 2254(98.7) | 4(0.2) | 15(0.6) | 12(0.5) |
| Ever been attacked with a knife/gun or other weapon by your husband/partner | 2258(98.8) | 6(0.3) | 17(0.8) | 3(0.1) |
| **Sexual Violence** |  |  |  |  |
| Ever been physically forced into unwanted sex by your husband/partner | 2136(93.5) | 35(1.5) | 101(4.4) | 13(0.6) |
| Ever been forced into other unwanted sexual acts by your husband/partner | 2233(97.7) | 14(0.6) | 32(1.4) | 6(0.3) |
| Ever been physically forced to perform sexual acts respondent didn't want to | 2206(96.5) | 17(0.8) | 52(2.3) | 9(0.4) |

**Figure 1.**
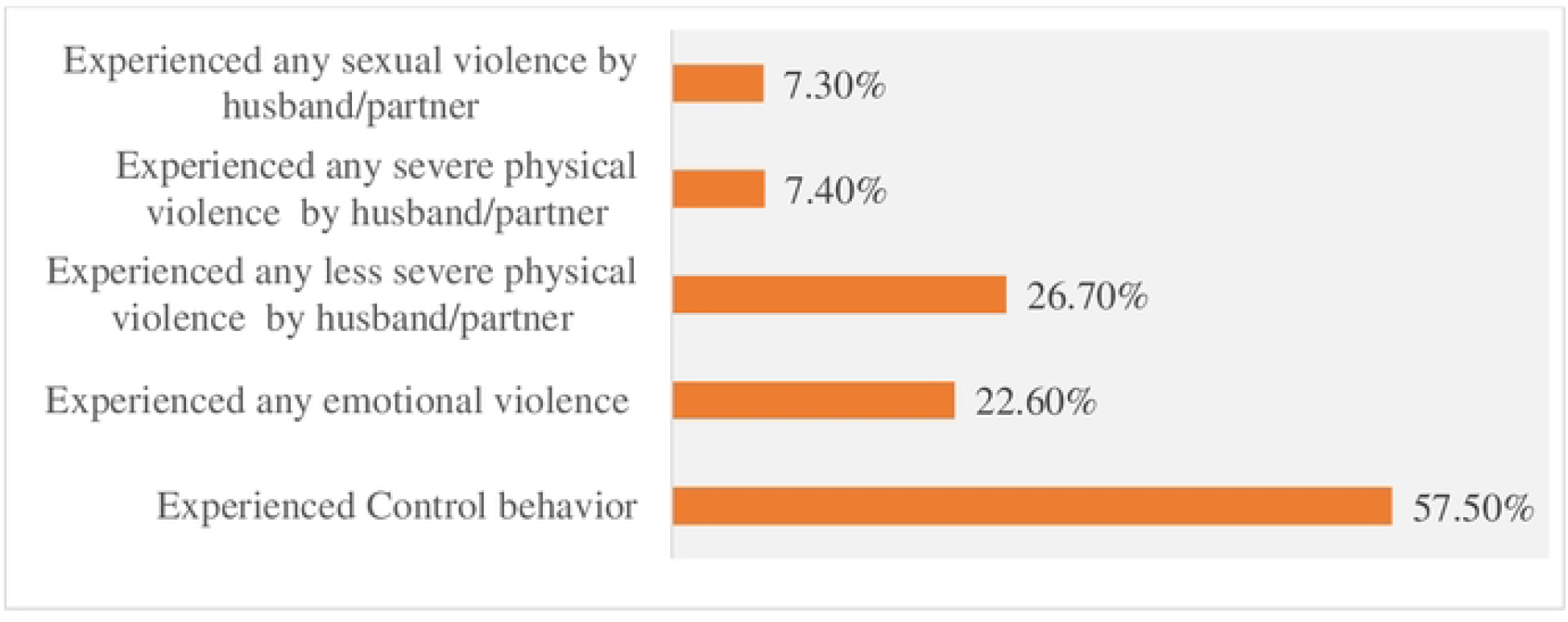
Intimate partner violence among women who gave birth in the last two years

### Magnitude of IPV

Out of the 2,284 study participants, 787 (34.5%) reported having experienced at least one form of IPV, while 1,497 (65.5%) indicated no history of experiencing any form of IPV (Figure 2)

**Figure 2.**
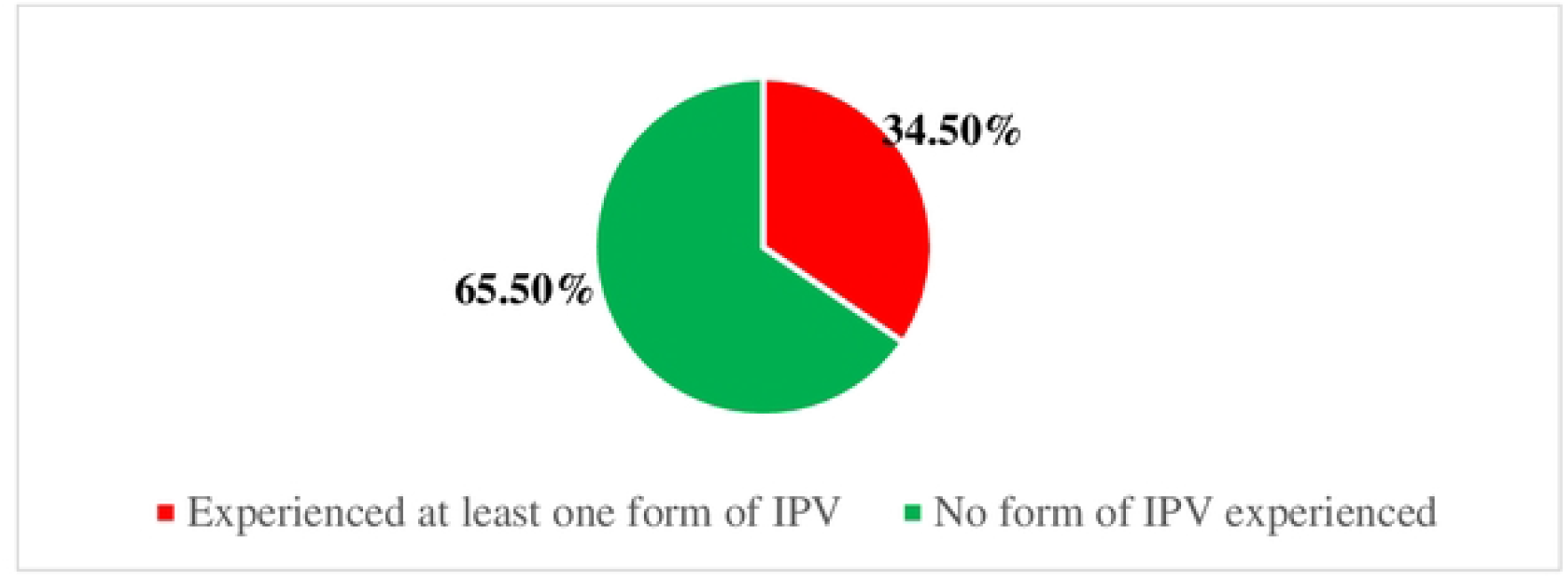
The magnitude of IPV among women of reproductive age who gave birth within two years preceding the survey

### Non-facility birth among women who were selected for intimate partners’ violence survey

Among women who gave birth in the last two years and were selected for the intimate partner violence survey, 438 (19.2%) had non-facility births, while 1,846 (80.8%) delivered in health facilities (Figure 3).

**Figure 3.**
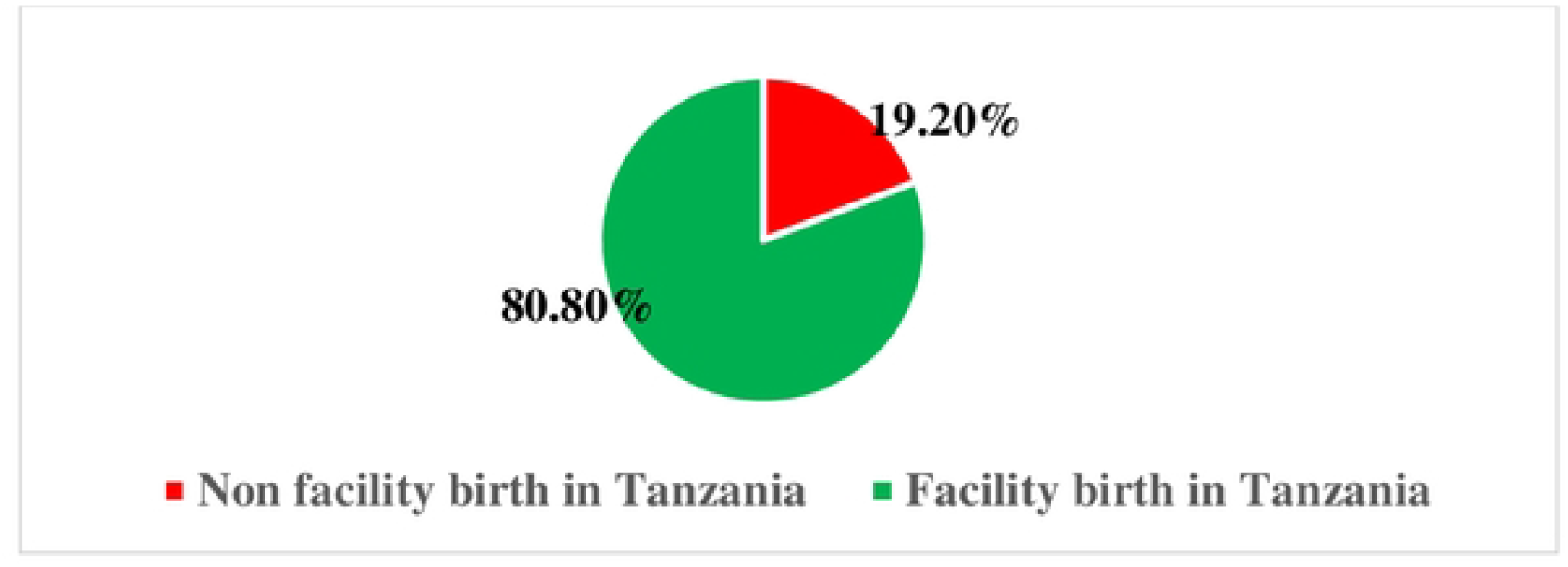
the proportion of non-facility births among women who gave birth two years preceding the survey and were included in the IPV survey questions

### The relationship between women’s characteristics and non-facility birth

Variables significantly associated with non-facility births included type of residence (X^2^=98.946, p<0.001), the highest education level (X^2^=130.981, p<0.001), wealth index (X^2^=223.370, p<0.001), number of ANC visits (X^2^=66.487, p<0.001), parity (X^2^=68.654, p<0.001), control behavior (X^2^=5.405, p=0.02), experiencing any emotional violence (X^2^=9.185, p=0.002), experiencing less severe violence by husband/partner (X^2^=10.975, p=0.001), experiencing severe violence by husband/partner (X^2^=9.059, p=0.003), and experiencing sexual violence by husband/partner (X^2^=9.641, p=0.002) (see Table 3).

**Table 3:** The relationship between women’s characteristics and place of childbirth.

| Variables | Place of Childbirth |  | X<sup>2</sup> | p-value |
| --- | --- | --- | --- | --- |
|  | Facility n(%) | Non-facility n(%) |  |  |
| **Experienced IPV** |  |  | 12.081 | 0.001 |
| No | 1241(82.9) | 256(17.1) |  |  |
| Yes | 605(76.9) | 182(23.1) |  |  |
| **Age Categories** |  |  | 3.053 | 0.217 |
| 15-19 years | 118(84.3) | 22(15.7) |  |  |
| 20-34 years | 1323(81.2) | 306(18.8) |  |  |
| 35-49 years | 405(78.5) | 110(21.5) |  |  |
| **Type of place of residence** |  |  | 98.946 | <0.001 |
| Urban | 563(94.6) | 32(5.4) |  |  |
| Rural | 1282(75.9) | 406(24.1) |  |  |
| **Highest educational level** |  |  | 130.981 | <0.001 |
| No education | 311(65.1) | 167(34.9) |  |  |
| Primary | 1092(81.8) | 243(6.1) |  |  |
| Secondary | 428(93.9) | 28(6.1) |  |  |
| Higher | 15(100) | 0(0.0) |  |  |
| **Wealth index** |  |  | 223.370 | <0.001 |
| Poorest | 320(61.5) | 200(38.5) |  |  |
| Poorer | 344(76.4) | 106(23.6) |  |  |
| Middle | 369(83.9) | 71(16.1) |  |  |
| Richer | 451(90) | 50(10) |  |  |
| Richest | 361(97) | 11(3) |  |  |
| **At least four ANC visits** |  |  | 66.487 | <0.001 |
| Yes | 1260(85.8) | 208(14.2) |  |  |
| No | 586(71.8) | 230(28.2) |  |  |
| **Parity of participants** |  |  | 68.654 | <0.001 |
| Para 0-1 | 397(92.1) | 34(7.9) |  |  |
| Para 2-4 | 1032(81.3) | 238(18.7) |  |  |
| Para 5-17 | 418(71.5) | 167(28.5) |  |  |

**Table 4:**
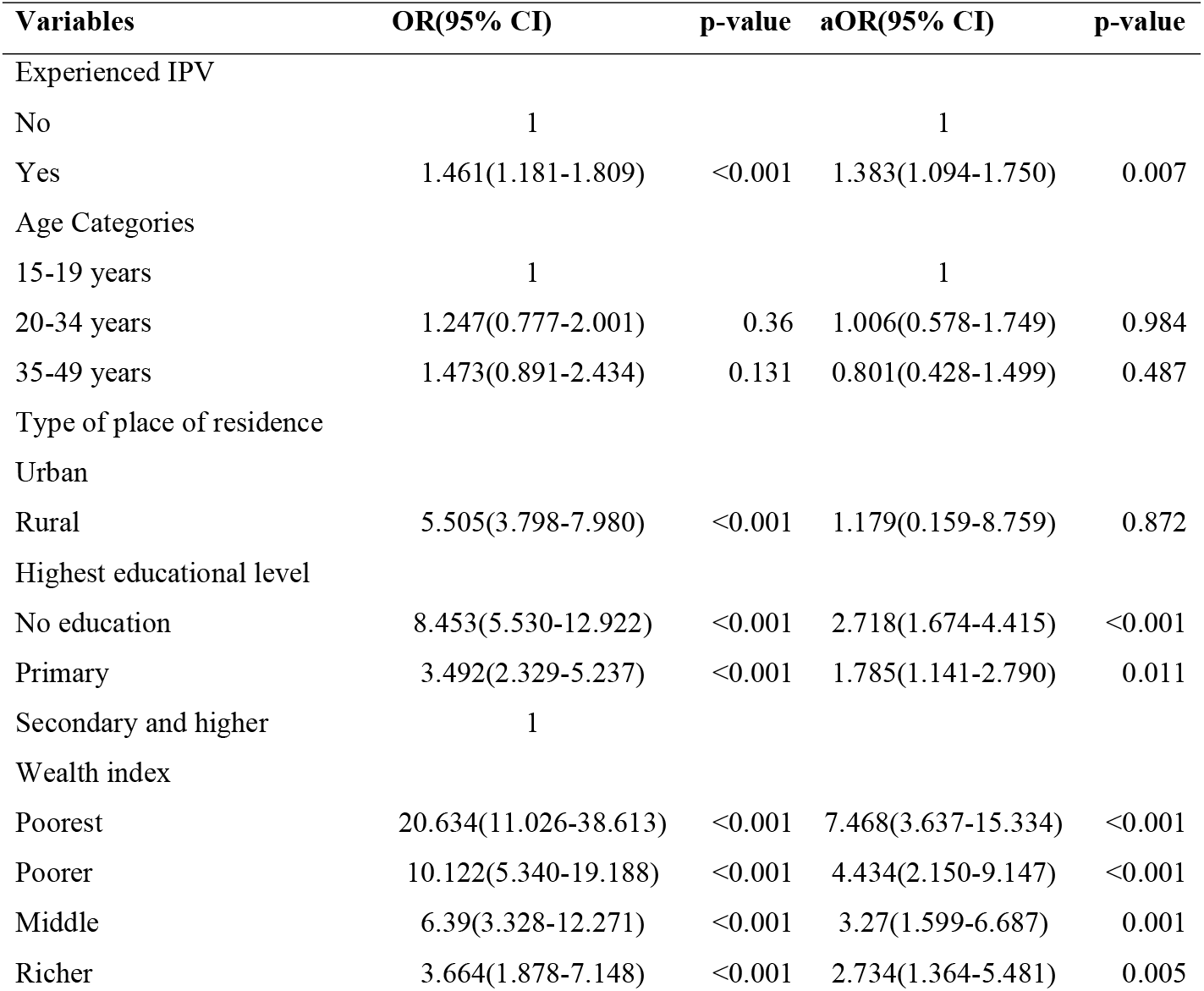
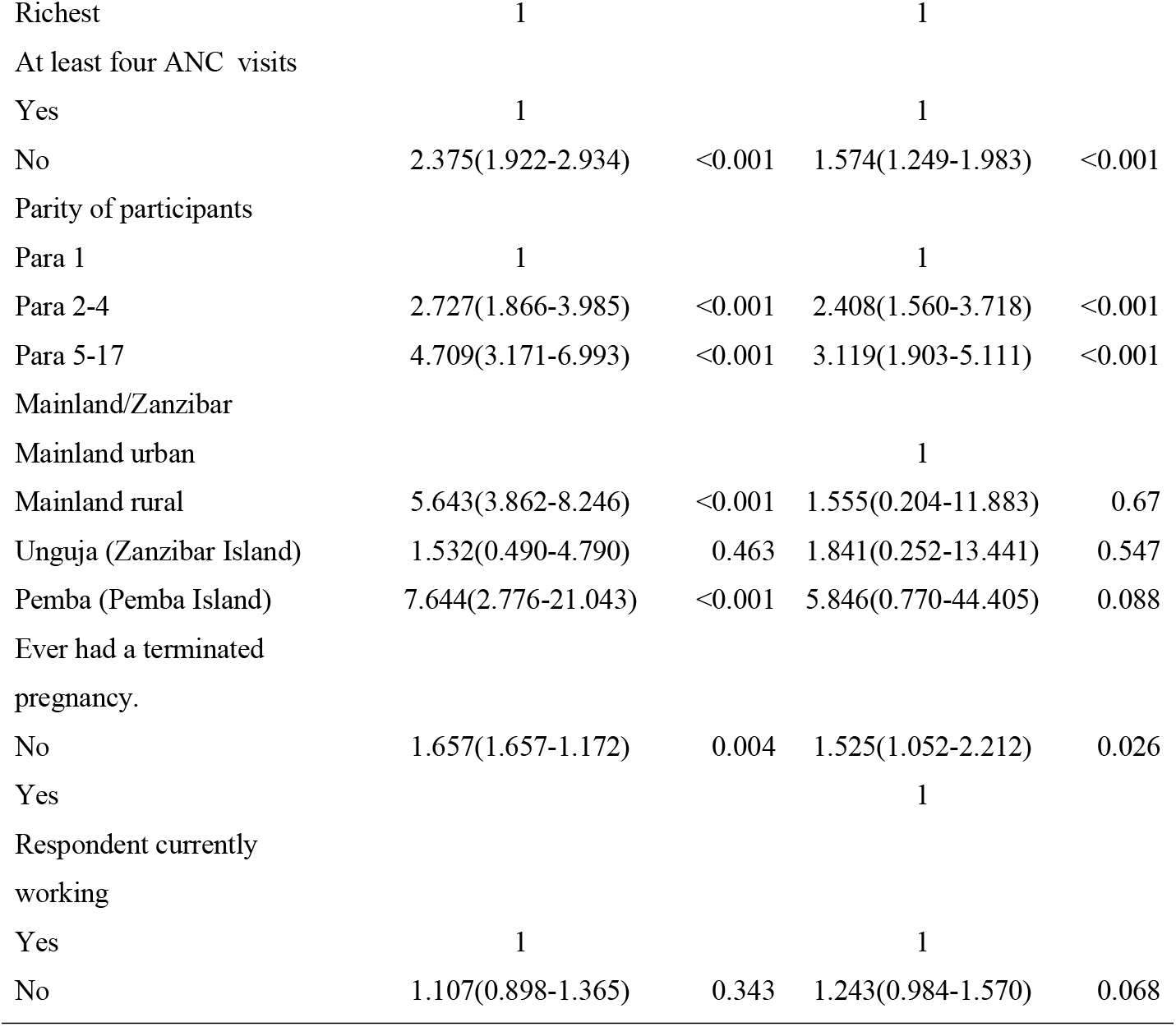
Predictors of Non-facility Birth with Unskilled Attendants.

| Variables | OR(95% CI) | p-value | aOR(95% CI) | p-value |
| --- | --- | --- | --- | --- |
| Experienced IPV |  |  |  |  |
| No | 1 |  | 1 |  |
| Yes | 1.461(1.181-1.809) | <0.001 | 1.383(1.094-1.750) | 0.007 |
| Age Categories |  |  |  |  |
| 15-19 years | 1 |  | 1 |  |
| 20-34 years | 1.247(0.777-2.001) | 0.36 | 1.006(0.578-1.749) | 0.984 |
| 35-49 years | 1.473(0.891-2.434) | 0.131 | 0.801(0.428-1.499) | 0.487 |
| Type of place of residence |  |  |  |  |
| Urban |  |  |  |  |
| Rural | 5.505(3.798-7.980) | <0.001 | 1.179(0.159-8.759) | 0.872 |
| Highest educational level |  |  |  |  |
| No education | 8.453(5.530-12.922) | <0.001 | 2.718(1.674-4.415) | <0.001 |
| Primary | 3.492(2.329-5.237) | <0.001 | 1.785(1.141-2.790) | 0.011 |
| Secondary and higher | 1 |  |  |  |
| Wealth index |  |  |  |  |
| Poorest | 20.634(11.026-38.613) | <0.001 | 7.468(3.637-15.334) | <0.001 |
| Poorer | 10.122(5.340-19.188) | <0.001 | 4.434(2.150-9.147) | <0.001 |
| Middle | 6.39(3.328-12.271) | <0.001 | 3.27(1.599-6.687) | 0.001 |
| Richer | 3.664(1.878-7.148) | <0.001 | 2.734(1.364-5.481) | 0.005 |
| Richest | 1 |  | 1 |  |
| At least four ANC visits |  |  |  |  |
| Yes | 1 |  | 1 |  |
| No | 2.375(1.922-2.934) | <0.001 | 1.574(1.249-1.983) | <0.001 |
| Parity of participants |  |  |  |  |
| Para 1 | 1 |  | 1 |  |
| Para 2-4 | 2.727(1.866-3.985) | <0.001 | 2.408(1.560-3.718) | <0.001 |
| Para 5-17 | 4.709(3.171-6.993) | <0.001 | 3.119(1.903-5.111) | <0.001 |
| Mainland/Zanzibar |  |  |  |  |
| Mainland urban |  |  | 1 |  |
| Mainland rural | 5.643(3.862-8.246) | <0.001 | 1.555(0.204-11.883) | 0.67 |
| Unguja (Zanzibar Island) | 1.532(0.490-4.790) | 0.463 | 1.841(0.252-13.441) | 0.547 |
| Pemba (Pemba Island) | 7.644(2.776-21.043) | <0.001 | 5.846(0.770-44.405) | 0.088 |
| Ever had a terminated pregnancy. |  |  |  |  |
| No | 1.657(1.657-1.172) | 0.004 | 1.525(1.052-2.212) | 0.026 |
| Yes |  |  | 1 |  |
| Respondent currently working |  |  |  |  |
| Yes | 1 |  | 1 |  |
| No | 1.107(0.898-1.365) | 0.343 | 1.243(0.984-1.570) | 0.068 |

### Predictors of Non-facility Birth attended by unskilled birth attendants

After controlling for confounders, the predictors of non-facility birth were ever experienced IPV (aOR=1.383, CI=1.094-1.750, p=0.007); level of education [No formal education (aOR=2.718, CI=1.674-4.415, p<0.001), primary education (aOR=1.785, CI=1.141-2.790, p=0.011)]; wealth index [poorest (aOR=7.468, CI=3.637-15.334, p<0.001), poorer (aOR=4.434 CI=2.150-9.147, p<0.001), middle (aOR=3.27, CI=1.599-6.687, p=0.001) and richer (aOR=2.734, CI=1.364-5.481, p=0.005)]; ANC attendance less than four visits (aOR=1.574, CI=1.249-1.983, p<0.001); Number of children ever born [Two to four (aOR=2.408, CI=1.560-3.718, p<0.001), more than four (aOR=3.119, CI=1.903-5.111, p<0.001)]; and ever terminated pregnancy (aOR=1.525, CI=1.052-2.212, p=0.026)

## Discussion

Results of this study demonstrate a significant prevalence of non-facility births of 19.2%, among women who took part in IPV interviews. The study also found that 35.5% of Tanzanian women had experienced intimate partner violence. In Tanzania, a history of intimate partner violence, low maternal education, low household economic status, fewer than four ANC visits, multiparity, and a history of pregnancy termination were all important predictors of non-facility childbirth.

With an alarming 19.2% prevalence of non-facility births among women who took part in IPV interviews in Tanzania, the study’s findings highlight a serious lack of access to maternal healthcare for victims of IPV. This noteworthy rate highlights the multiple vulnerabilities that these women encounter, such as logistical, emotional, and physical obstacles that prevent them from seeking essential healthcare during pregnancy and childbirth (7). Similarly, a previous study conducted in sub-Saharan Africa revealed that 36.1% of all births were attended by unskilled birth attendants (8). Among women who did not receive care from a skilled birth attendant (SBA), 17.6% were assisted by traditional birth attendants (TBAs), 12.3% relied on a friend or relative, and 4.9% gave birth without any assistance (8).

Even after adjusting for potential confounders, the results show that IPV is a significant predictor of non-facility births attended by unskilled birth attendants. Interestingly, compared to women who had never experienced IPV, those who had experienced IPV were 1.38 times more likely to give birth outside of a medical facility. Due to fear, limited mobility, and a lack of support from their partners, women who experience intimate partner violence frequently have limited access to maternal healthcare services, according to studies conducted in sub-Saharan Africa (9–11). Women’s autonomy can be increased and facility-based deliveries can be encouraged by addressing IPV through integrated maternal health programs that include IPV screening and support services. These findings collectively emphasize the urgent need for targeted interventions to address the intersection of IPV and maternal health, ensuring that no woman is forced to give birth outside a safe and supportive healthcare environment due to violence or fear.

The study also found a strong and significant association between non-facility birth and maternal education level. Women with only a primary education had nearly twice the chance of giving birth outside of a medical facility, while those without any formal education were nearly three times more likely to do so. Previous studies have demonstrated that women with only a primary education or no formal education have lower health literacy, less knowledge of the benefits of maternal health, and less self-efficacy when navigating healthcare systems(12). Similar findings have been reported in Tanzania and other low-resource settings where maternal health use is closely linked to women’s educational attainment (13–15). Informed decision-making and more facility births can result from increasing maternal health literacy and women’s educational opportunities.

The study also found that a strong predictor of non-facility birth was the household wealth index. As poverty increased, the likelihood of giving birth outside of a medical facility increased as well.

The likelihood of a non-facility birth was seven times higher for women from the poorest households than for those from the wealthiest. This is in line with previous studies that show poverty is associated with cost-related barriers like lack of household financial decision-making authority, indirect care costs, and transportation challenges(16,17). To assist low-income women in overcoming financial barriers, social protection programs such as maternal health vouchers, conditional cash transfers, and community transportation initiatives should be extended(18).

The study also found that women who had fewer than four ANC visits were almost twice as likely to give birth outside of a hospital as women who had four or more visits. Given the strong link between lower ANC attendance and non-facility births, the WHO recommends that at least four ANC visits are necessary for early detection and treatment of complications. (19). Data from past studies carried out in sub-Saharan Africa supports this conclusion, demonstrating that ANC visits provide an opportunity to educate and encourage facility-based deliveries (20–22). Attendance may be increased and the number of institutional births may rise if ANC services are made available, and comprehensive, and encourage facility delivery through community-based education and mobile ANC services.

The study also discovered that women who had multiple children were more likely to choose non-facility birth. Compared to primigravida women, those who had two or four children were twice as likely to give birth outside of a hospital. Non-facility childbirth was three times more common among those with five or more children. Similar findings have been reported in previous studies conducted in sub-Saharan Africa Individualized birth preparedness planning and additional family planning counseling should be provided to women who have given birth more than once to address concerns and encourage facility-based deliveries.

This study has several shortcomings despite its insightful findings. First, it is more difficult to prove a link between intimate partner violence and non-facility births due to the cross-sectional design. Future research should use longitudinal designs to better understand causal relationships over time to mitigate this. Second, self-reported information on IPV and place of birth could be underreported due to social desirability bias and recall issues. Researchers recommend the use of mixed-method techniques, such as triangulation with medical facility records and qualitative interviews, to validate results. Third, the study’s focus on Tanzania restricts the findings’ applicability to other contexts with distinct sociocultural and medical backgrounds. Comparative studies and research expansion across various geographic areas can improve the results’ applicability and guide more comprehensive maternal health interventions.

## Conclusion

According to this study, there are significant gaps in maternal healthcare access, and specific interventions are needed to address the systemic issues causing non-facility births. For women, especially those impacted by intimate partner violence, socioeconomic inequalities, low health literacy, and insufficient prenatal care pose serious challenges. A multifaceted strategy is needed to address these issues, which includes strengthening ANC outreach programs, increasing educational and financial assistance programs, and incorporating IPV support into maternal health services. Healthcare systems in Tanzania can improve facility-based delivery rates, increase maternal autonomy, and lower maternal and neonatal risks by giving priority to these strategies.

## Abbreviations

ANC: Antenatal Clinic
IPV: Intimate Partner Violence
PHC: Population and Housing Census
SBA: Skilled Birth Attendant
SDG: Sustainable Development Goal
SPSS: Statistical Package for Social Sciences
TBA: Traditional Birth Attendant
TDHS-MIS: Tanzania Demographic and Health Survey and Malaria Indicator Survey
WHO: World Health Organization

## Acknowledgments

The dataset was made available by MEASURE DHS, for which the authors are extremely grateful. Our sincere gratitude is also extended to the Institute of Science Tokyo for kindly hosting the research fellowship, the Matsumae International Foundation for their generous sponsorship, and the University of Dodoma for providing a study leave that made this study possible.

## Copyrighted figures

No figure requires a copyright

## Dual publication

No part of this paper has been published

## Data availability statement

The data set is available on the following website https://dhsprogram.com/

## Authors Contribution

FVM did the conceptualization process, wrote the methodology section, obtained access to the dataset, carried out the data analysis, and was in charge of writing the manuscript. KN oversaw data extraction, directed the methodology’s development, gave thorough critical feedback on the manuscript, and offered expert supervision throughout the conceptualization phase. By offering thorough criticism and perceptive edits, YT, AM, HS, and MO significantly contributed. The final version was unanimously approved by all authors, who actively participated in the manuscript review process.

